# Reflux Esophagitis is Associated with Higher Risks of Acute Stroke and Transient Ischemic Attacks in Patients Hospitalized with Atrial Fibrillation: A Nationwide Inpatient Sample Analysis

**DOI:** 10.1101/2020.08.07.20169482

**Authors:** Yi Jiang, Konstantinos Damiris, Giselle A. Suero-Abreu, Sushil Ahlawat

## Abstract

**Objective:** Reflux esophagitis (RE) is a subset of gastroesophageal reflux disease (GERD) with endoscopic evidence of esophageal inflammation, which has been linked to an increased incidence of atrial fibrillation (AF). However, data on the effect of RE on patient outcomes is limited. We sought to examine the potential association of RE with outcomes of patients with AF in a nationwide study.

**Methods:** The National Inpatient Sample (NIS) database was queried to identify hospitalized adult patients with AF and RE between 2010 and 2014. Primary outcomes included inpatient mortality, length of stay (LOS), and total hospital charges. AF related complications such as acute stroke, transient ischemic attack (TIA) and acute heart failure were assessed as secondary outcomes. Propensity score matching and multivariate regression analysis were used.

**Results:** 667,520 patients were admitted for primary diagnosis of AF out of which 5,396 had a secondary diagnosis of RE. In the AF with RE cohort, the average age was 73.6 years, 41.5% were male, and 79.9% were Caucasian. There was a greater prevalence of concomitant dyslipidemia, chronic liver disease and chronic pulmonary disease (p <0.01) when compared to the AF without RE cohort. Patients with AF and RE also had higher incidence of acute strokes and TIAs (p<0.05), longer LOS (p<0.001), and higher hospital charges (p<0.05) with no difference in acute heart failure (p=0.08), hospital mortality (p=0.12), or CHA_2_DS_2_-VAS score (p=0.67).

**Conclusion:** In hospitalized patients with AF, RE was associated with a higher rate of acute stroke and TIA, longer LOS, and greater hospital charges.

## Introduction

Gastroesophageal reflux disease (GERD) is a pathologic condition that develops from the reflux of stomach contents, which causes a variety of troublesome clinical symptoms with or without complications [1]. With an estimated prevalence of 8-33% worldwide, and 18-28% in the United States (US) alone; it carries a cost of more than 9-10 billion dollars per year in the US [2-4]. Notably, GERD is the most common indication for esophagogastroduodenoscopy in the US [5]. Reflux esophagitis (RE) describes a subset of patients with GERD who have endoscopic evidence of esophageal inflammation.

Atrial fibrillation (AF) is the most common cardiac rhythm disorder [6], affecting 2.3 million individuals in the US [7]. Its effects are profound, as it is associated with a 3-5 fold increase for stroke [8] and a two fold increase of all-cause mortality [9]. CHA2DS2-VASc risk stratification score has been used widely for estimation of stroke risk for non-valvular AF in adults [10] and can assist with the decision of initiating anticoagulation therapy [11].

The coexistence of GERD/RE and AF, is frequently encountered in the clinical setting and increases the complexity of patients’ diagnosis and treatment. Pathogenetically, AF has been shown to be associated with inflammatory disorders and abnormalities of left atrium geometry [12]. Based on the close proximity of the esophagus and the left atrium, the interaction between GERD/RE and AF has been proposed. GERD was reported to be associated with an increased risk of AF [13-15] in some studies. However, the results have been controversial due to limited sample sizes or underpowered study design. Moreover, the impact of GERD on outcomes of AF related hospitalization has been underexplored.

Bunch et al [16] suggested that the presence of esophagitis instead of GERD symptoms alone plays a pivotal role in triggering and promoting AF. Therefore, we aimed to examine the influence of RE, instead of the clinical diagnosis of GERD, on the outcomes in adult patients hospitalized for AF.

## Materials and Methods

### 2.1. Data Source and Study Population

In this study the National Inpatient Sample (NIS) database [17] was utilized to assess patient demographics, hospital characteristics and inpatient admission outcomes of 20% of all in-patient hospitalizations within the United States. Patient data was obtained using the International Classification of Diseases-Ninth Edition Revision, Clinical Modification (ICD-9 CM) codes allowing for identification of diagnoses and procedures associated with patients’ hospitalization. This retrospective cohort study examined adult patients, ages 18-90 years, admitted between 2010 and 2014. Primary diagnosis (first or second admission diagnosis) of AF (ICD-9 code 427.31) and secondary diagnosis of RE (ICD-9 code 530.11) was utilized. Patients with AF and co-existing RE were compared to those without RE. Collected data included patient demographics, comorbidities, post-hospitalization disposition, and other various outcomes (Tables 1-3). Patient medical comorbidities were categorized using the approved Elixhauser Comorbidity Index (ECI) score [18], which uses 29 common medical conditions and proceeds to calculate a weighted compiled score used to predict hospital resource use and mortality amongst hospitalized patients. Primary outcomes included inpatient mortality, length of stay (LOS), and accrued total hospital charges. Secondary outcomes included AF with and without RE leading to complications including acute stroke, TIA and acute heart failure.

**Table 1.**
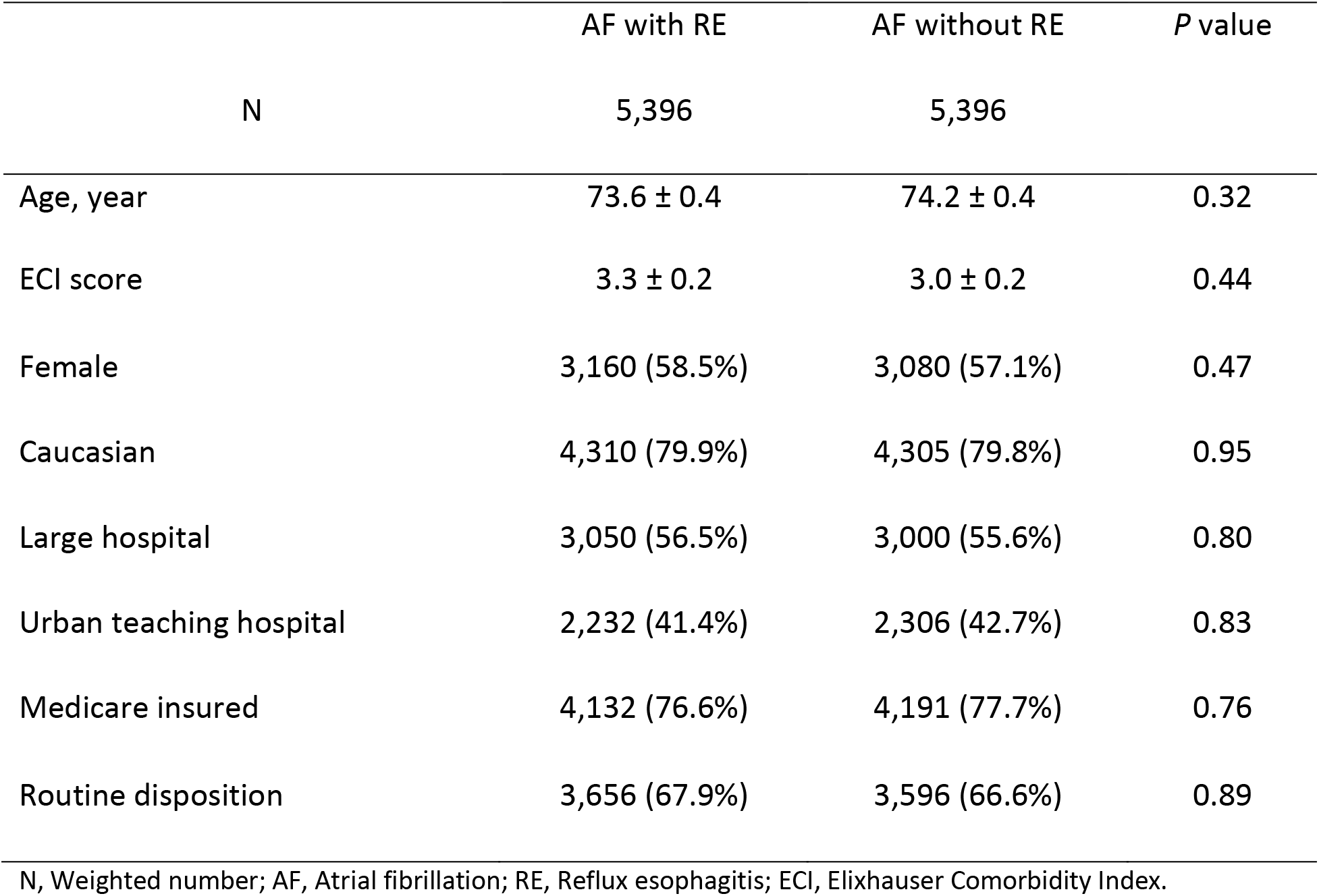
Demographics and baseline characteristics of patients admitted for atrial fibrillation with and without reflux esophagitis.

**Table 2.** Comparison of selected comorbidities of patients admitted for atrial fibrillation with and without reflux esophagitis.

|  | AF with RE | AF without RE | P value |
| --- | --- | --- | --- |
| N | 5,396 | 5,396 |  |
| CHA<sub>2</sub>DS<sub>2</sub>-VASc score |  |  | 0.67 |
| 0 | 139 (2.6%) | 129 (2.4%) |  |
| 1 | 407 (7.5%) | 357 (6.6%) |  |
| >=2 | 4,850 (89.9%) | 4,910 (91.0%) |  |
| Tobacco use disorder | 1,359 (25.2%) | 1,364 (25.3%) | 0.96 |
| Dyslipidemia | 2,743 (50.8%) | 2,386 (44.2%) | 0.002 |
| Obesity | 744 (13.8%) | 694 (12.9%) | 0.53 |
| Congestive heart failure | 169 (3.1%) | 159 (2.9%) | 0.80 |
| Hypertension | 3,839 (71.1%) | 3,819 (70.8%) | 0.85 |
| Diabetes mellitus | 1,428 (26.5%) | 1,468 (27.2%) | 0.69 |
| History of stroke, TIA, or thromboembolism | 560 (10.4%) | 526 (9.7%) | 0.62 |
| Thromboembolic disease | 40 (0.7%) | 15 (0.3%) | 0.13 |
| Acute myocardial infarction | 20 (0.4%) | 45 (0.8%) | 0.18 |
| Prior myocardial infarction | 432 (8.0%) | 392 (7.3%) | 0.52 |
| Prior PCI | 417 (7.7%) | 422 (7.8%) | 0.94 |
| Prior CABG | 387 (7.2%) | 382 (7.1%) | 0.93 |
| Valvular disease | 184 (3.4%) | 159 (2.9%) | 0.55 |
| Prior valvular surgery | 134 (2.5%) | 188 (3.5%) | 0.17 |
| Cardiogenic shock | 5 (0.1%) | 20 (0.4%) | 0.18 |
| Cardiac arrest | 15 (0.3%) | 10 (0.2%) | 0.65 |
| Chronic liver disease | 144 (2.7%) | 60 (1.1%) | 0.0067 |
| Chronic pulmonary disease | 1,543 (28.6%) | 1,146 (21.2%) | 0.0001 |
| Renal failure | 694 (12.9%) | 600 (11.1%) | 0.21 |
| Catheter ablation | 119 (2.2%) | 188 (3.5%) | 0.08 |
| Open surgical ablation | 20 (0.4%) | 10 (0.2%) | 0.41 |
| Electric cardioversion | 446 (8.3%) | 471 (8.7%) | 0.69 |

AF, Atrial fibrillation; RE, Reflux esophagitis; TIA, Transient ischemic attack; PCI, Percutaneous coronary intervention; CABG, Coronary artery bypass graft surgery.

**Table 3.** Multivariate regression analysis of inpatient outcomes for patients admitted for atrial fibrillation with and without reflux esophagitis

|  | AF with RE | AF without RE | Unadjusted odds ratio or coefficient (95% CI) | *P* value | Adjusted odds ratio or coefficient* (95% CI) | *P* value |
| --- | --- | --- | --- | --- | --- | --- |
| Acute stroke | 25 | 10 | 2.51 | 0.015 | 2.34 | 0.028 |
| and TIA | (0.46%) | (0.19%) | (1.2, 5.24) |  | (1.1, 4.99) |  |
| Acute heart failure | 109 | 129 | 0.84 | 0.194 | 0.79 | 0.076 |
|  | (2.02%) | (2.39%) | (0.65, 1.09) |  | (0.61, 1.02) |  |
| Hospital mortality | 64 | 50 | 1.3 | 0.161 | 1.35 | 0.123 |
|  | (1.19%) | (0.93%) | (0.9, 1.89) |  | (0.92, 1.99) |  |
| LOS, day | 4.05 | 3.48 | 0.56 | <0.0001 | 0.54 | 0.0001 |
|  | (0.11) | (0.10) | (0.28, 0.85) |  | (0.27, 0.82) |  |
| Total |  |  |  |  |  |  |
| hospitalization | 36,095 | 32,114 | 3,980 | 0.0240 | 3,890 | 0.0250 |
| charges, dollars | (1,409) | (1,122) | (518, 7,443) |  | (483, 7,297) |  |

Data are presented as mean (standard errors) or absolute numbers (%)
AF, Atrial fibrillation; RE, Reflux esophagitis; CI, Confidence interval; TIA, Transient ischemic attack; LOS, Length of stay.
\*Adjusted for age, sex, race, primary insurance payer, hospital type, hospital bed size, income quartile, CHA<sub>2</sub>DS<sub>2</sub>-VASC score, Elixhauser Comorbidity Index score, dyslipidemia, chronic liver disease, and chronic pulmonary disease.

### 2.2. Statistical Analysis

All statistical analyses were performed using SAS Survey Procedures (SAS 9.4, SAS Institute Inc, Cary, NC, USA). Confounding variables included patient age, sex, race, primary insurance payer, hospital type, hospital bed size, hospital region and hospital teaching status. These variables were controlled using propensity score matching [19,20] with use of multivariate logistic regression model. With use of 8-to-1-digit match, each admission of AF with RE was matched with one admission from AF without RE. National estimates were calculated after accounting for the sample design elements (clusters, strata, and trend weights) provided by the NIS. All continuous variables are reported as weighted means ± standard errors (SE). All categorical variables are displayed as weighted numbers (N) and percentages (%). Paired t-tests were used for the comparison of normally distributed continuous variables, while Wilcoxon Rank-Sum Test was used for non-normally distributed continuous variables. Categorical variables were analyzed using the Rao-Scott modified chi-square test. Multivariate linear regression was used to estimate the average change in LOS and total hospital charges after adjusting for patient demographics (age, sex, race), hospital bed size, insurance type, median household income, ECI score, CHA_2_DS_2_-VASC Score, dyslipidemia, chronic liver disease and chronic pulmonary disease. Multivariate logistic regression calculated the odds ratio of outcomes after adjusting for the prior mentioned confounding variables.

### 2.3. Ethical information

The NIS database is publicly available and includes only de-identified patient demographics. This study was a retrospective study; therefore, no patients were actively involved, making Institutional Review Board approval non-applicable.

## Results

### 3.1. Patient Demographics and Baseline Characteristics

A total of 667,520 patients were admitted for primary diagnosis of AF, of which 5,396 patients had documented diagnosis of coexisting RE. These patients were matched with 5,396 AF patients without RE. The AF with RE cohort in this study was predominantly a geriatric Caucasian population with an average age of 73.6, and an average ECI score of 3.3. 58.5% of patients were female. 76.6% patients were insured by Medicare, and 67.9% were discharged routinely (Table 1). Compared to AF patients without RE, there were no differences in the demographic variables examined. The AF with RE cohort was associated with significant comorbid conditions. About 90% patients in this cohort had a CHA_2_DS_2_-VASc Score ≥ 2, 71.1% had coexisting hypertension, 26.5% had diabetes, and half of the patients had dyslipidemia. AF with RE patients had a significantly greater prevalence of concomitant dyslipidemia (p = 0.0018), chronic liver disease (p = 0.0067) and chronic pulmonary disease (p = 0.0001) compared to AF without RE (Table 2). No statistically significant differences in ECI score, CHA_2_DS_2_-VASc Score or AF-related procedure rates were found between AF with RE vs. AF without RE group.

### 3.2. Outcomes and Regression Analysis of AF patients with and without RE

When compared to AF without RE group, AF with RE patients had a higher incidence of acute strokes and TIAs [0.46% vs. 0.19%, adjusted odds ratio (aOR) = 2.34, 95% confidence interval (CI) 1.1-4.99, p < 0.05), longer LOS (4.05 days vs 3.48 days, adjusted coefficient 0.54, 95% CI 0.27-0.82, p = 0.0001), and higher hospital charges ($36,095 vs. $32,114, adjusted coefficient 3,890, 95% CI 483-7,297, p < 0.05). There were no differences in either acute heart failure or hospital mortality (Table 3).

## Discussion

This nationwide study examined the impact of RE on the outcomes of patients hospitalized for AF. Our study population was predominantly geriatric Caucasian patients who had high comorbidity burden and high risk for stroke based on the CHA_2_DS_2_-VASc score. We suspect that elderly patients who have a high comorbidity burden may be indicated for endoscopy more often secondary to alarm symptoms and complicated GERD [21]. They may also be indicated for hospitalization to treat active AF-related comorbidities more often than the general population. This study demonstrated that AF with RE was independently associated with an increased risk of inpatient acute stroke and TIAs, longer LOS, and higher hospitalization costs compared to AF without RE.

Several epidemiological studies have suggested that GERD, particularly esophagitis, is associated with increased risk of AF onset and maintenance [13,14]. Some studies further demonstrated that proton-pump inhibitors help ameliorate AF symptoms and facilitate conversion from AF to sinus rhythm in a subset of patients with GERD [22,23], which indirectly suggests the relationship between GERD and AF. The potential GERD-associated arrhythmogenic mechanisms have been proposed based on three main factors from animal models and clinical findings [14,24]: esophageal inflammation, autonomic nerve activation, and mechanical irritation from the esophagus to the nearby left atrium.

Despite both AF and RE being common diseases with significant causal relationship, little is known regarding how RE affects the outcomes of AF.

RE potentially contributes to an increased stroke risk in AF by both the localized and systemic inflammation present with the disease. Importantly, it has been suggested in both human and animal models that RE develops as a cytokine-mediated inflammatory injury [25] and not as a caustic chemical injury, as traditionally theorized. Souza et al [26] developed a RE rat model after esophagoduodenostomy and found the inflammation did not start in the mucosa, but deep in the epithelium by histological study. Later it was found that stopping proton-pump inhibitors in patients with severe RE was associated with esophageal inflammation without loss of surface cells [27,28]. It was demonstrated that esophageal epithelial cells secreted interleukin (IL)-8, IL-1β, and other potent proinflammatory cytokines when exposed to acidic bile salts [26]. Additionally, it was reported that inflammatory pathways are not only involved in the initiation and maintenance of AF, but also contribute to both electrical and structural atrial remodeling and thrombogenesis in patients with AF [29]. Many systemic inflammatory diseases are accompanied by adverse atrial remodeling and an enhanced risk of stroke [30,31]. Systemic inflammatory disorders can cause inflammatory injury to the coronary microcirculation, leading the microvascular dysfunction as well as myocardial fibrosis [12]. The extension of the systemic inflammatory process to the atrial wall has been evidenced by significant left atrial abnormalities on cardiac images [32]. Moreover, states of inflammation cause deranged adipogenesis of epicardium, which is connected with the myocardium through an unobstructed microcirculation. Dysfunctional epicardial adipose tissue expands its mass and secretes proinflammatory adipocytokines, which are further linked to the anatomical and pathophysiological substrates for AF [33,34] and its severity [35]. In conjunction, RE associated inflammatory injury can potentially lead to microvascular and microcirculation changes in the cardiovascular system from cytokine release, leading to dysregulated electrical activity. Secondarily, the close proximity of the esophagus and the left atrium can lead to extension of inflammation to the atrial wall and epicardium, further influencing the aforementioned left atrial geometric change and epicardial adipose tissue expansion. These heart structural abnormalities, together with reinforced inflammatory status, subsequently trigger the pathogenesis of thromboembolic events.

Reactive oxygen species (ROS) and oxidant stress [36] are also related to RE pathogenesis and affect the risk of AF and related adverse events. Feagins et al [37] has reported that upon exposure to acid and bile salts, cultured esophageal squamous cells increase ROS production. The ROS, together with hypoxia from tissue inflammation, induced the production of hypoxia-inducible factors (HIF)-2 α, which mediate the expression of pro-inflammatory molecules [38] and function as initiators of the cytokine-mediated inflammation. ROS and oxidant stress have been recognized as important contributors to atrial remodeling in AF [39-41]. Studies have shown that several ion channels and their regulators expressed in the atria are sensitive to redox state under oxidant stress [42]. A decreased atrial calcium current and a diminished contractile response to adrenergic agonists have been observed during experimental inhibition of glutathione synthesis [43]. It was proposed that pathologic processes that increase oxidant production via any of the several pathways might have a similar electrical and contractile phenotype that is associated with AF [39]. Hence, ROS production in RE exacerbates both inflammatory and oxidative pathways, which have been implicated in the AF-related atrial geometry abnormalities and further thromboembolic events.

Interestingly, despite the statistically significant higher rate of inpatient stroke and TIAs in the AF with RE group, there was no difference in overall CHA_2_DS_2_-VASc score or any of its components. RE increased acute stroke and TIA risk in a way that exceeds that predicted by the presence of traditional cardiovascular risk factors. Similarly, it is reported that the rate of stroke in patients with known AF and systemic autoimmune diseases is greater than can be explained by the CHA_2_DS_2_-VASc score [44,45]. Notably, clinical studies have reported that the risk of AF and related thromboembolic events are particularly apparent in clinically severe inflammatory disease [31] Anatomically, pericardial fat volume expansion, which is linked to AF onset and adverse events, has also been proved to be proportional to the inflammatory disease clinical severity and the intensity of inflammation [12]. Collectively, thromboembolic risk of AF may be positively correlated with the severity and intensity of local or systemic inflammation. The CHA_2_DS_2_-VASc score does not incorporate measures of inflammation or direct assessments of the atrial myopathy or abnormal geometry, therefore the predictive accuracy in severe inflammatory disease is compromised [12]. Indeed, some have proposed that the CHA_2_DS_2_-VASc score be modified in patients who have a systemic inflammatory disorder [46].

This study showed higher rates of chronic pulmonary and liver diseases in hospitalized AF patients with RE compared with those without RE. These patients with RE may be symptomatic from diseases of other systems and may seek medical attention more often than those without reflux symptoms. Repeat medical attention may result in identification of disease from other systems like chronic pulmonary or liver diseases, as demonstrated in this study. More investigation will be needed to uncover the underlying correlation between RE and pulmonary or liver disease in AF patients.

Our study has several strengths. To the best of our knowledge, this is a leading nationwide study to address the impact of RE on outcomes for hospitalized AF patients. Our study went beyond the correlation studies of these two diseases and focused on the clinical outcomes. The results suggested that additional information about severity and intensity of inflammation may help improve the predictive accuracy of thromboembolic risk score in AF patients with significant inflammatory disease. Also, we used propensity score matching and multivariate analysis to isolate the effect of RE after adjusting for a full list of possible confounders including ECI score and CHA_2_DS_2_-VASc score. On the other hand, limitations of this study are worth noting. Because of inherent limitations from NIS, all diagnoses included depend on the accuracy of ICD-9 codes and medical documentation. The supportive clinical symptoms, signs, diagnostic images, labs, and medication administration were shared between physicians and coding professionals, but are not included in the NIS database. The quality control is regularly performed by the third parties: The Agency for Healthcare Research and Quality (AHRQ) and Healthcare Cost and Utilization Project (HCUP).

In conclusion, this nationwide study examined the impact of RE on the outcomes of patients hospitalized for AF. The results suggested RE independently increased stroke and TIA risk in hospitalized patients with AF. This finding may guide clinical decision making on anticoagulation and/or other treatment modalities targeted at alleviating cardiac arrhythmia. Patients with significant inflammatory diseases with or without left atrium geometry changes may benefit from anticoagulation for stroke risk reduction even with low CHA_2_DS_2_-VASc scores. In the future, prospective studies with long term follow up, focusing on assessing the effect of inflammation severity and intensity on thromboembolic risk in AF patients with GERD related diseases, may help better risk stratification. Understanding the complex pathophysiological process of RE-associated AF might help to identify specific anti-reflux or anti-inflammatory strategies for the treatment of AF and the prevention of AF-related adverse events [47].

## Data Availability

Data related to this manuscript was obtained from the National Inpatient Database which could be obtained through HCUP - AHRQ

## Author Contributions and Notes

Y.J and K.D. designed research, Y.J and K.D. performed research, Y.J analyzed data; and Y.J and K.D. and G.A.S.A. wrote the paper. Y.J and K.D. and G.A.S.A and S.A critical review of paper.

The authors declare no conflict of interest.

## Acknowledgments

We thank Luka Petrovic, MD, Cardiology Fellow at Mount Sinai Morningside Health Care System for his professional suggestions from Cardiology perspectives.

